# Performance of ChatGPT, GPT-4, and Google Bard on a Neurosurgery Oral Boards Preparation Question Bank

**DOI:** 10.1101/2023.04.06.23288265

**Authors:** Rohaid Ali, Oliver Y. Tang, Ian D. Connolly, Jared S. Fridley, John H. Shin, Patricia L. Zadnik Sullivan, Deus Cielo, Adetokunbo A. Oyelese, Curtis E. Doberstein, Albert E. Telfeian, Ziya L. Gokaslan, Wael F. Asaad

**Affiliations:** Department of Neurosurgery, The Warren Alpert Medical School of Brown University, Providence, RI, USA; Department of Neurosurgery, University of Pittsburgh, Pittsburgh, PA, USA; Department of Neurosurgery, Massachusetts General Hospital, Boston, MA, USA

**Keywords:** Neurosurgery, medical education, surgical education, residency education, artificial intelligence, large language models, ChatGPT

## Abstract

**Background:** General large language models (LLMs), such as ChatGPT (GPT-3.5), have demonstrated capability to pass multiple-choice medical board examinations. However, comparative accuracy of different LLMs and LLM performance on assessments of predominantly higher-order management questions is poorly understood.

**Objective:** To assess performance of three LLMs (GPT-3.5, GPT-4, and Google Bard) on a question bank designed specifically for neurosurgery oral boards examination preparation.

**Methods:** The 149-question Self-Assessment Neurosurgery Exam (SANS) Indications Exam was used to query LLM accuracy. Questions were input in a single best answer, multiple-choice format. Chi-squared, Fisher’s exact, and univariable logistic regression tests assessed differences in performance by question characteristics.

**Results:** On a question bank with predominantly higher-order questions (85.2%), ChatGPT (GPT-3.5) and GPT-4 answered 62.4% (95% confidence interval [CI]: 54.1-70.1%) and 82.6% (95% CI: 75.2-88.1%) of questions correctly, respectively. In contrast, Bard scored 44.2% (66/149, 95% CI: 36.2-52.6%). GPT-3.5 and GPT-4 demonstrated significantly higher scores than Bard (both *P*<0.01), and GPT-4 significantly outperformed GPT-3.5 (*P*=0.023). Among six subspecialties, GPT-4 had significantly higher accuracy in the Spine category relative to GPT-3.5 and in four categories relative to Bard (all *P*<0.01). Incorporation of higher-order problem solving was associated with lower question accuracy for GPT-3.5 (OR=0.80, *P*=0.042) and Bard (OR=0.76, *P*=0.014), but not GPT-4 (OR=0.86, *P*=0.085). GPT-4’s performance on imaging-related questions surpassed GPT-3.5’s (68.6% vs. 47.1%, *P*=0.044) and was comparable to Bard’s (68.6% vs. 66.7%, *P*=1.000). However, GPT-4 demonstrated significantly lower rates of “hallucination” on imaging-related questions than both GPT-3.5 (2.3% vs. 57.1%, *P*<0.001) and Bard (2.3% vs. 27.3%, *P*=0.002). Lack of question text description for imaging predicted significantly higher odds of hallucination for GPT-3.5 (OR=1.45, *P*=0.012) and Bard (OR=2.09, *P*<0.001).

**Conclusion:** On a question bank of predominantly higher-order management case scenarios intended for neurosurgery oral boards preparation, GPT-4 achieved a score of 82.6%, outperforming ChatGPT and Google’s Bard.

## Introduction

Growing interest has surrounded the ability of artificial intelligence (AI) to guide clinical decision-making and care, especially given recent documentation of the ability of general Large Language Models (LLMs) like ChatGPT (OpenAI; San Francisco, CA) to pass graduate-level and certification examinations in fields including medicine,^1^ law,^2^ and business.^3^ In a prior analysis, we found ChatGPT achieved a passing score of 73.4% on a 500-question module emulating the neurosurgery written board examinations, with lower accuracy on questions that were lengthier, incorporated higher-order problem-solving, or involved imaging.^4^ While ChatGPT has been available for public use since November 2022 as a “GPT-3.5” release, OpenAI released an updated model, GPT-4, on March 14, 2023. Like its predecessor, GPT-4 was trained using both supervised and unsupervised learning techniques on a large corpus of Internet text data, followed by fine-tuning via reinforcement learning with human feedback. GPT-4 has achieved passing scores in over 25 standardized examinations, including scoring in the 90^th^ percentile of a simulated bar exam, compared to GPT-3.5 scoring in the 10^th^ percentile.^5^ Evidence of performance improvements of over 20% on all three United States Medical Licensing Exam (USMLE) exams has also been documented.^6^ GPT-4 additionally has introduced multimodal capabilities, including the ability to evaluate image inputs, that have yet to be released for public use.

In response to the popularity of ChatGPT and GPT-4, various leading software companies have introduced their own language models, showcasing remarkable advancements in artificial intelligence. One such example is the Bard chatbot, developed by Google’s parent company, Alphabet Inc. (Mountain View, California). Launching on March 21, 2023, Bard has garnered considerable attention as Google’s foray into the chatbot domain, sparking intriguing discussions about the future of search technology.

A key distinction between Bard and ChatGPT and GPT-4 lies in Bard’s ability to access and incorporate information from the internet in real-time when generating responses. This contrasts with ChatGPT and GPT-4, which rely on prior training data up until September 2021 and do do not have webcrawling capabilities incorporated presently. Incorporating real-time web search capabilities, Bard, in theory, could offer users more current and contextually pertinent information. However, direct comparisons between the two models are only just beginning to be undertaken. Notably, there have been no head-to-head comparisons of Bard and ChatGPT within the context of any clinical board examination.

The performance of LLMs like GPT-4 on open-ended oral medical examinations is less understood. In the setting of neurosurgery, the American Board of Neurological Surgery (ABNS) oral board examination is composed of three 45-minute sessions and most commonly taken 2-3 years after residency graduation, in contrast to the written board exam that is intended for earlier-stage trainees.^7^ The oral board examination is widely considered the more rigorous and difficult assessment. While the first-time pass rate for the American Board of Neurological Surgery written board examination has exceeded 96% since 2018, the pass rate for the oral board examination has ranged between 81-90% during the same period.^8^

The goals of this study were to **a)** assess the performance of three LLMs on a question bank with higher-order questions more representative of oral board topics and **b)** elucidate differences in accuracy and performance by question characteristics between LLMs.

## Methods

This study assessed the performance of three LLMs: ChatGPT, GPT-4, and Google Bard. Performance of LLMs on the neurosurgery oral board examination was proxied using the Self-Assessment Neurosurgery Exam (SANS) Indications Exam, a 149-question module designed specifically for oral boards preparation, focusing on surgical indications, diagnostic testing, and interventional decision-making. Notably, this module is written in a multiple-choice format, which differs from the open-ended approach of the oral boards. As described earlier,^4,9^ question characteristics including subspecialty, word length, and incorporation of higher-order problem-solving were independently collected by two authors (RA and OYT), with disagreements adjudicated by a third (IDC). All classification was blinded, without investigator knowledge of any LLM’s answers to the questions.

Questions were assessed in a single best answer multiple-choice format, with the question stem reproduced verbatim (**Figure 1**). Because multimodal input has yet to be incorporated into any of the three LLMs, questions with imaging data had only the text portion of the question stem input for evaluation. Questions that an LLM opted not to answer, such as due to citing insufficient contextual data, were classified as incorrect. Additionally, for questions with imaging, we tracked responses with confabulations or “hallucinations,” which were defined as scenarios where an LLM asserted inaccurate facts or contextual data that it falsely believed were correct in its answer. This phenomenon has been well documented by OpenAI among ChatGPT and, to a lesser degree, GPT-4.^5^ Data were collected on 3/12/23 for ChatGPT, 3/14/23 (day of release) for GPT-4, and 4/1/23 for Google Bard.

**Figure 1:**
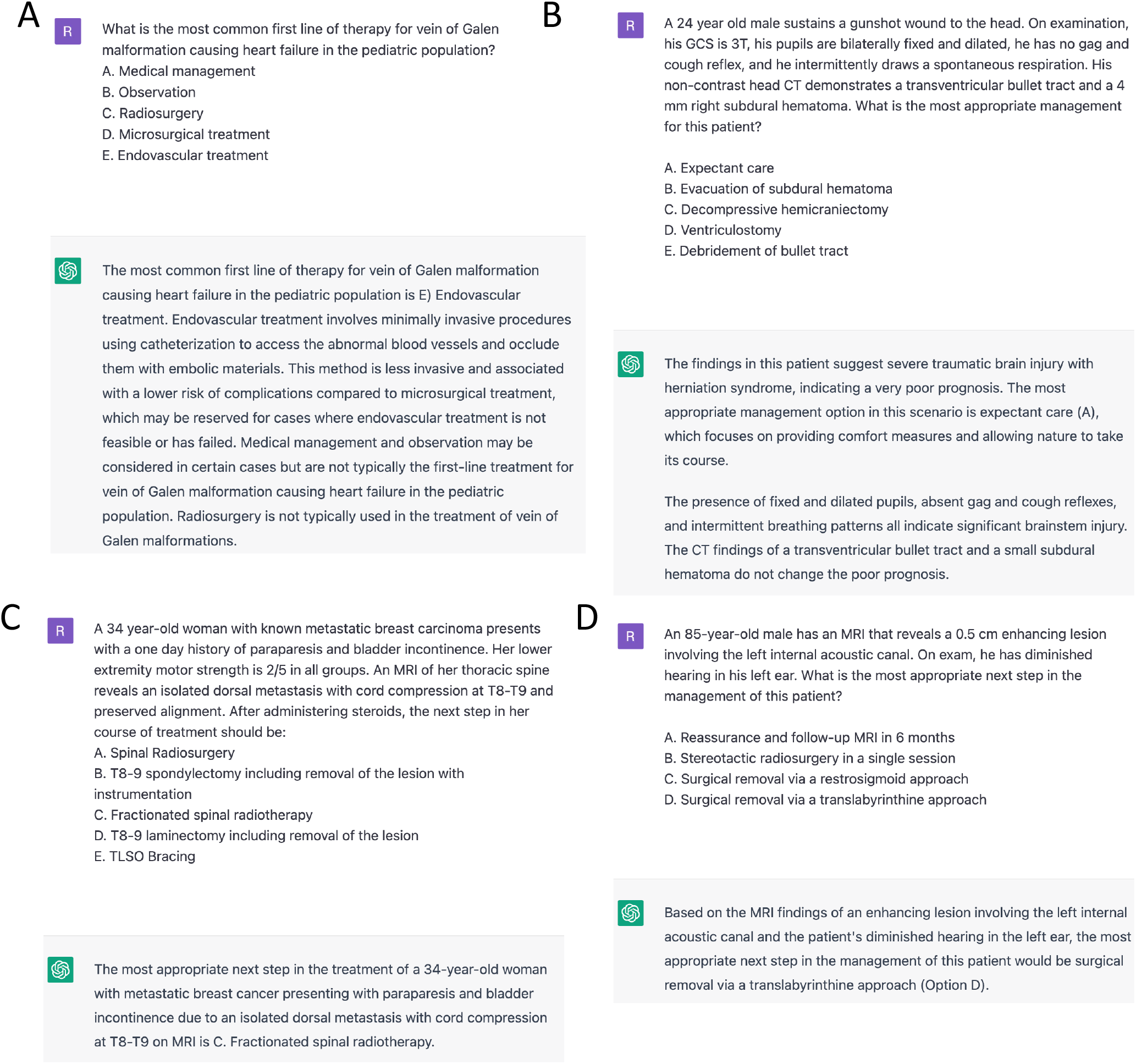
Representative Indications Question Bank Examination Responses by ChatGPT (GPT-3.5) and GPT-4. Images are attributed to ChatGPT (OpenAI; San Francisco, CA) and GPT-4 (OpenAI; San Francisco, CA), which can be found at h ttps://chat.openai.com/. **A:** First-order question on the most common first-line treatment option for a vein of Galen malformation causing heart failure. **B:** Higher-order question on expectant management for a severe traumatic brain injury patient with a poor prognosis. Higher-order questions were classified as those involving multiple problem-solving steps to arrive at the correct answer, such as identifying an unknown diagnosis followed by choosing the best course of management (as opposed to just identifying the diagnosis in a first-order question). Notably, in its answer rationale, ChatGPT demonstrated an ability to declare futility. **C:** A higher-order question on management of breast cancer spinal metastases answered incorrectly by both GPT-3.5 and GPT-4. In this scenario, ChatGPT recommended fractionated radiotherapy for a non-radiosensitive lesion causing myelopathy. **D:** A higher-order question on management of presumed vestibular schwannoma in a frail octogenarian answered incorrectly by both GPT-3.5 and GPT-4.

All analyses and visualizations were performed using R Version 4.1.2 (Foundation for Statistical Computing, Vienna, Austria) and the *matplotlib* package on Python (Python Software Foundation, Wilmington, DE), respectively. Associations between category-level performance were queried using linear regression. Differences in performance were assessed using chi-squared, Fisher’s exact, and univariable logistic regression tests. For all analyses, statistical significance was assessed at *P*<0.05. This study was conducted in accordance with Strengthening the Reporting of Observational Studies in Epidemiology (STROBE) reporting guidelines.

## Results

### Performance of ChatGPT, GPT-4, and Bard on Indications Question Bank

On the 149-question question bank focused on oral boards topics, GPT-3.5 (ChatGPT) and GPT-4 achieved scores of 62.4% (93/149, 95% confidence interval [CI]: 54.1-70.1%) and 82.6% (123/149, 95% CI: 75.2-88.1%), respectively (**Table 1**). GPT-3.5 performed significantly worse on the oral boards question bank, relative to GPT-3.5’s performance on a previously reported 500-question written boards question bank (62.4% vs. 73.4%, *P*=0.013). GPT-4’s 82.6% score surpassed GPT-3.5’s scores on both the written (*P*<0.001) and oral boards (*P*=0.023) question banks (**Figure 2** and **Figure 3**). GPT-4 correctly answered all 93 questions that GPT-3.5 did and demonstrated significantly better performance in the Spine subspecialty, specifically (90.5% vs. 64.3%, *P*=0.008).

**Table 1:** Performance of ChatGPT (GPT-3.5), GPT-4, and Google Bard on Oral Board Examination-Focused Question Bank.

| Question Category | Questions | GPT-3.5 Performance | GPT-4 Performance | Google Bard Performance | *P*-Value |  |  |
| --- | --- | --- | --- | --- | --- | --- | --- |
|  |  |  |  |  | 3.5 vs. 4 | 3.5 vs. Bard | 4 vs. Bard |
| Overall | 149 | 93/149 (62.4%) | 123/149 (82.6%) | 66/149 (44.2%) | <0.001 | 0.002 | <0.001 |
| General | 33/149 (22.1%) | 20/33 (60.6%) | 27/33 (81.8%) | 11/33 (33.3%) | 0.102 | 0.048 | <0.001 |
| Functional | 8/149 (5.4%) | 7/8 (87.5%) | 7/8 (87.5%) | 5/8 (62.5%) | 1.000 | 0.569 | 0.569 |
| Peripheral Nerve | 20/149 (13.4%) | 11/20 (55.0%) | 15/20 (75.0%) | 4/20 (20.0%) | 0.320 | 0.048 | 0.001 |
| Spine | 42/149 (28.2%) | 27/42 (64.3%) | 38/42 (90.5%) | 27/42 (64.3%) | 0.008 | 1.000 | 0.008 |
| Tumor | 21/149 (14.1%) | 11/21 (52.4%) | 15/21 (71.4%) | 5/21 (23.8%) | 0.341 | 0.111 | 0.004 |
| Vascular | 25/149 (16.8%) | 17/25 (68.0%) | 21/25 (84.0%) | 14/25 (56.0%) | 0.321 | 0.561 | 0.062 |

Performance of ChatGPT on oral board examination-focused question bank. Overall performance and subspecialty breakdown are reported. Differences in performance between LLMs were queried using chi-squared and Fisher's exact tests.

**Figure 2:**
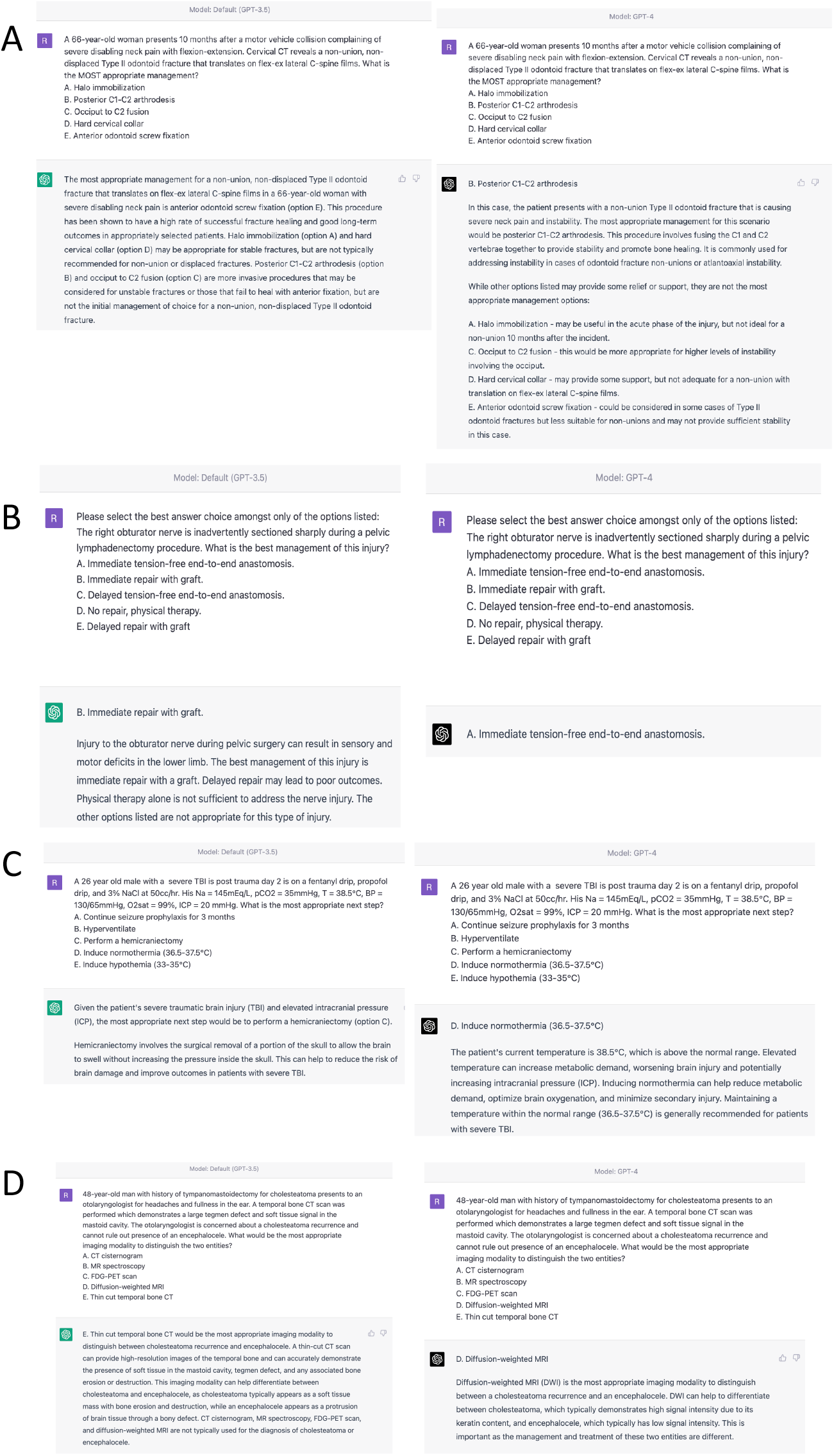
Contrast in Question Answers Between ChatGPT (GPT-3.5) and GPT-4. Four representative questions across different subspecialties answered incorrectly by ChatGPT (GPT-3.5) but correctly by GPT-4. Images are attributed to ChatGPT (OpenAI; San Francisco, CA) and GPT-4 (OpenAI; San Francisco, CA), which can be found at h ttps://chat.openai.com/. **A:** Question on management of a type II odontoid fracture. In this case, GPT-4 correctly recognized that anterior odontoid screw fixation was a less appropriate treatment modality for a chronic fracture exhibiting non-union, compared to posterior C1-C2 fusion. **B:** Question on management of an intraoperative transection of the obturator nerve. **C:** Question on management of a severe traumatic brain injury patient with elevated intracranial pressure. In this case, only GPT-4 recognized the patient’s hyperthermia and recommended inducing normothermia as the next best step of management. **D:** Question on best imaging modality to differentiate cholesteatoma recurrence and encephalocele.

**Figure 3:**
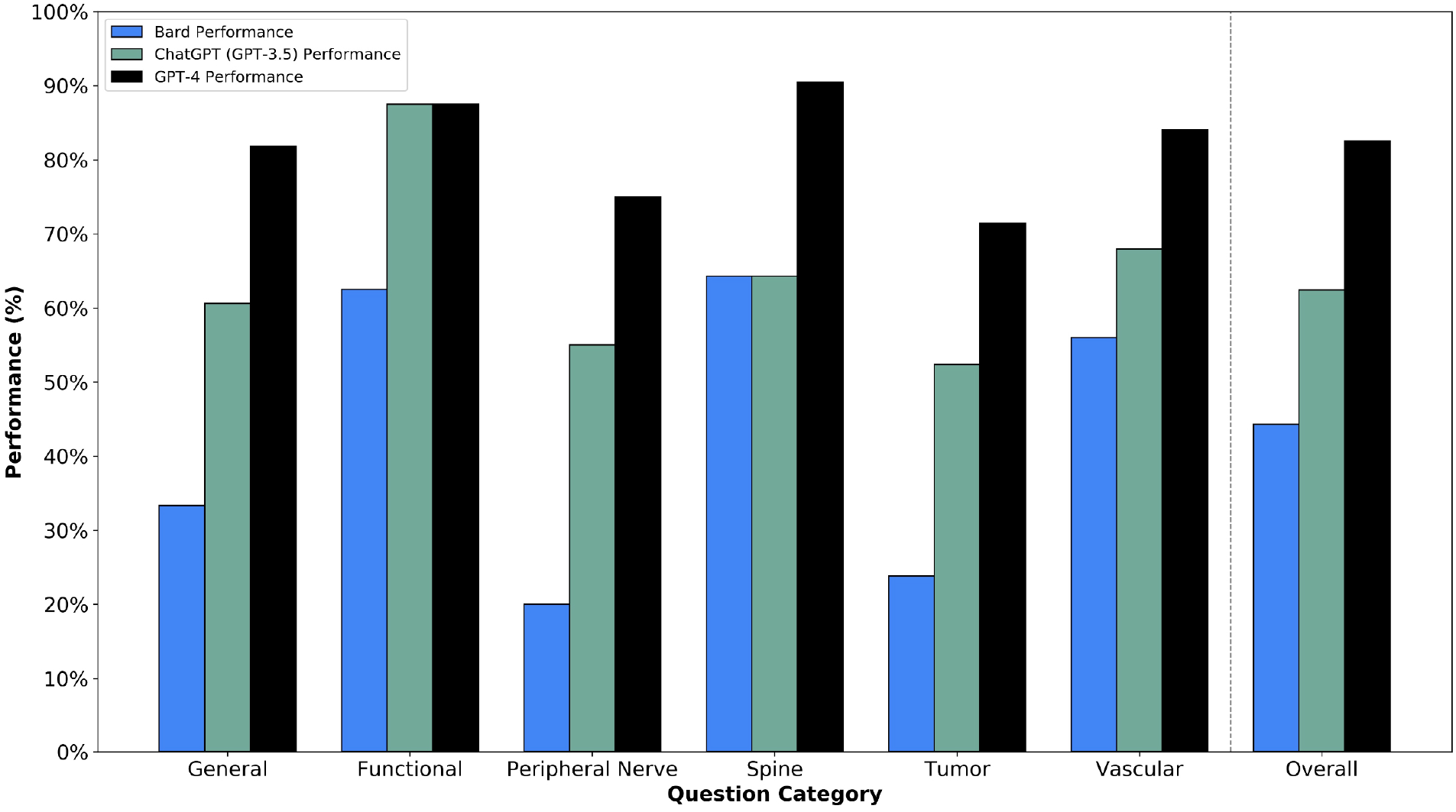
Performance of ChatGPT (GPT-3.5), GPT-4, and Bard on the Indications Question Bank. Histograms of performance of ChatGPT (GPT-3.5; white), GPT-4 (gray), and Bard (red) on the Indications question bank, including overall and subspecialty-level performance. Error bars denote 95% confidence interval for scores.

In contrast, Bard provided correct answers for only 44.2% (66/149, 95% CI: 36.2-52.6%) of questions, returning incorrect answers for 45.0% (67/149) and opting entirely out of answering 10.7% (16/149) questions. Of the 16 questions Bard declined to answer, 2 involved imaging, while 14 were solely text-based (**Figure 4A-C**). There were no instances of GPT-3.5 or GPT-4 declining to answer a solely text-based question. Both GPT-3.5 (62.4% vs. 44.2%, *P*=0.002) and GPT-4 (82.6% vs. 44.2%, *P*<0.001) had superior performance on the Indications Exam, relative to Bard. Bard had significantly lower scores on the General and Peripheral Nerve categories compared to both GPT-3.5 (both *P*<0.05) and GPT-4 (both *P*<0.002). Additionally, GPT-4 alone outperformed Bard on Spine (*P*<0.008) and Tumor (*P*=0.004) questions. Of the 66 questions answered correctly by Bard, GPT-3.5 and GPT-4 responded correctly on 77.3% (51/66) and 97.0% (64/66), respectively.

**Figure 4:**
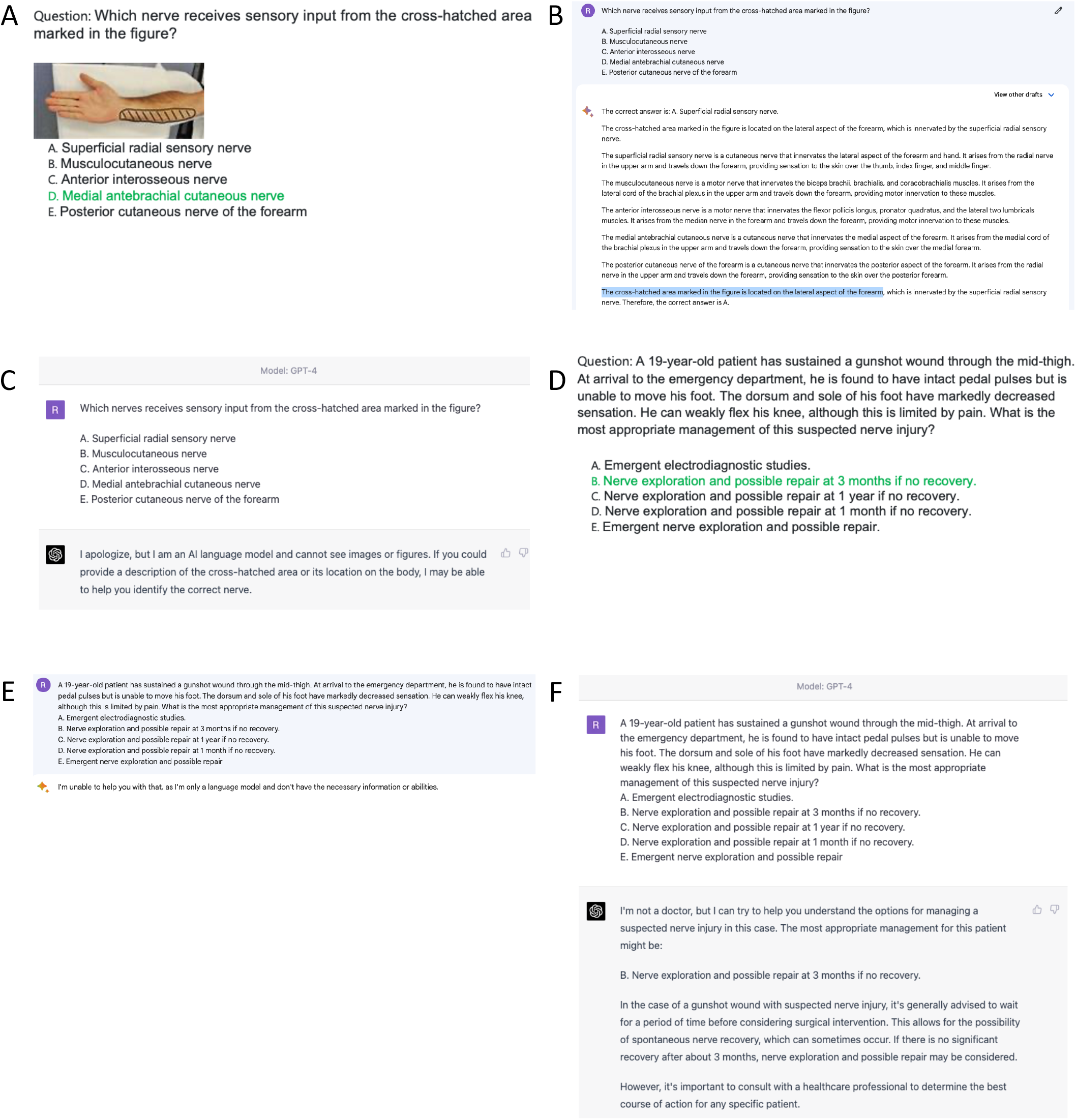
Differences in Performance Between Bard and GPT-4. Representative questions from the Indications Exam demonstrating notable differences in performance between Bard and GPT-4. Images are attributed to the Self-Assessment Neurosurgery Exam (SANS) Indications Exam (Congress of Neurological Surgeons; Schaumburg, IL), Bard (Alphabet Inc.; Mountain View, CA), and GPT-4 (OpenAI; San Francisco, CA). **A:** Question on management of a peripheral nerve injury due to gunshot wound. This was a text-based question with no related imaging. **B:** Screenshot of Bard’s response, which deferred on answering the question due to not having the “necessary information or abilities.” This scenario happened for 14 solely text-based questions in the examination for Bard but did not occur a single time for GPT-4. **C:** Screenshot of GPT-4’s response, where the correct course of management was selected. **D:** Question on identifying the nerve that supplies sensory input to the medial forearm. An image labeling the medial forearm was included in the question stem but could not be input into either Bard or GPT-4 due to present lack of multimodal capability. **E:** Screenshot of Bard’s response, where the labeled part of the imaging is incorrectly inferred as the lateral forearm (highlighted). This is an example of a “hallucination,” where GPT-4 cited a rationale for its answer explanation that could not be backed up by any of the input related to the question stem. **F:** GPT-4 declines to answer the question, due to lacking sufficient context.

### Question Characteristics and LLM Accuracy

Higher-order questions were significantly more represented in the Indications question bank, relative to the prior written boards question bank (85.2% vs. 7.4%, *P*<0.001). While higher-order problem-solving was predictive of lower question accuracy for GPT-3.5 (OR=0.80, *P*=0.042) and Bard (OR=0.76, *P*=0.014), this association was not significant for GPT-4 (OR=0.86, *P*=0.085). Notably, GPT-4 was able to answer higher-order questions involving challenging concepts like declaring medical futility (**Figure 1B**) but continued to struggle in other scenarios, such as incorporating disease-specific management considerations (**Figure 1C**) or factoring in patient-level characteristics like frailty (**Figure 1D**). Question length did not predict performance by GPT-3.5, GPT-4, or Bard.

### Performance on Imaging-Based Questions

51 questions (34.2%) incorporated imaging into the question stem. Due to multimodal input presently being unavailable for public use, only question text was provided to these models. Both GPT-3.5 and GPT-4 opted to answer 44 of these questions (86.3%), while declining to answer 7 (13.7%) due to insufficient context. In contrast, Bard returned an answer for 96.1% (49/51) of all imaging-based questions. GPT-4’s performance on imaging-related questions surpassed GPT-3.5’s (68.6% vs. 47.1%, *P*=0.044) and was comparable to Bard’s (68.6% vs. 66.7%, *P*=1.000).

On the 44 imaging-related questions attempted by GPT-3.5 and GPT-4, hallucinations were exhibited in 27.3% (12/44) and 2.3% (1/44) of answers, respectively. Bard demonstrated evidence of hallucinations in 57.1% (28/49) of these questions it attempted to answer, including 47.1% (8/17) of correct answers and 62.5% (20/32) of incorrect answers. For example, in questions where a specific area of the image is labeled, Bard would assert which portion of the image was labeled in its answer explanation, without any imaging input or context clues in the question stem text to inform this (**Figure 4D-F**). Both GPT-3.5 (27.3% vs. 57.1%, *P*=0.006) and GPT-4 (2.3% vs. 57.1%, *P*<0.001) had significantly lower rates of hallucination than Bard. Moreover, GPT-4 had a lower hallucination rate than GPT-3.5 (2.3% vs. 27.3%, *P*=0.002). A lack of text description for imaging in the question stem was associated with significantly higher likelihood of hallucination for GPT-3.5 (OR=1.45, *P*=0.012) and Bard (OR=2.09, *P*<0.001), but not GPT-4 (OR=1.03, *P*=0.547).

## Discussion

In summary, GPT-4 achieved a score of 82.6% on a question bank of predominantly higher-order diagnostic and management multiple-choice questions designed for oral boards preparation in the field of neurosurgery. This study represents the first of its kind to focus on predominantly higher-order questions in a medical subspecialty domain and the first in neurosurgery to compare performance of multiple LLMs. GPT-4 demonstrated improved performance in question categories for which GPT-3.5 exhibited lower accuracy, such as incorporating higher-order problem-solving or using context clues alone to answer imaging-related questions. In addition, the study revealed that GPT-4 outperformed Google Bard in all categories, underscoring the critical need for neurosurgeons to stay up-to-date on emerging LLMs and their varying levels of performance for potential application. GPT-3.5’s comparatively worse performance on the Indications question bank (62.4%) relative to its written boards results (73.4%) is possibly attributable to the greater representation of higher-order questions in the former.

As LLMs are increasingly challenged with more open-ended queries, the issues raised by a phenomenon known as “hallucinations” has become more pressing. Hallucinations describe instances in which LLMs assertively incorporate erroneous details into their generated responses. Notably, from a neurological viewpoint, such behavior might more properly be described as “confabulation.” This is particularly concerning in the context of neurosurgery, a high-stakes specialty where the consequences of hallucinations could potentially lead to catastrophic mistakes. Therefore, to develop trust in such systems, we must rigorously validate their performance on increasingly higher-order and open-ended scenarios. Recognizing the importance of addressing hallucinations, we developed methods to quantify them, which are essential to further understanding and to ensure that LLMs like GPT-4 can be safely and effectively integrated into clinical decision-making processes. Our findings demonstrate GPT-4’s improved capability to correctly answer higher-order management-focused evaluations of neurosurgical knowledge, reduced rates of hallucination, and an ability to navigate challenging topics like futility. However, the potential ethical and legal implications of using LLMs in clinical practice must be carefully considered.

There are additionally two notable changes to the neurosurgery written and oral boards examination process over the past decade, which warrant discussion in the context of the present study’s findings. First, the ABNS written (primary) exam has been intentionally modified over the past decade to serve as “mastery” assessments. In close collaboration with the American Association of Neurological Surgeons (AANS), Congress of Neurological Surgeons (CNS), and Society of Neurological Surgeons (SNS), considerable resources have been devoted to ensuring that the content tested on these examinations is as transparent and accessible as possible. This likely influences the availability of written exam content on the Internet, thereby providing an excellent foundation for training Large Language Models (LLMs). Similarly, it is also worth noting that the knowledge of oral examination content and commonly tested management concepts accessible online, even in a multiple-choice form like the SANS Indications Exam, may also enhance the training data available to LLMs. Nevertheless, it is conceivable that the successful performance of LLMs on these more open-access multiple-choice assessments may not fully translate to comparable performance when confronted with more unpredictable and unique real-world situations, which is more reflective of the oral boards testing environment and actual clinical practice. This potential gap in application may potentially already be appreciated by the poorer performance of GPT-3.5 and Bard on higher-order questions.

Second, the oral boards have undergone reorganization into a three-session format, including two sessions with standardized clinical scenarios with a wide range of acceptable responses. However, the third session is based on the candidate’s own cases as an attending and, despite the candidate being the definitive authority on the knowledge of how these cases were managed, is traditionally the most frequently failed session of the oral exam. This paradox demonstrates the importance of distinguishing possession of knowledge from application of knowledge, especially to complicated cases with individual-level considerations and significant equipoise, which compose a significant portion of neurosurgical practice. When considering the optimal approach for certification of future neurosurgeons, the utility of multiple-choice examinations, which can now be passed by LLMs, warrants further assessment. While performance on these tests serves as an indicator of possessing foundational knowledge, the oral boards examination highlights the significance of thoroughly probing broader management decisions in an open-ended and verbal format to determine if that knowledge can be applied appropriately, safely, and compassionately. AI programs will prove to be valuable resources, supplying clinicians with rapidly accessible and reliable information. However, it is the responsibility of the clinician to integrate this data with the unique circumstances of each patient. In other words, while AI algorithms may exhibit remarkable knowledge capabilities, it is ultimately the clinician who must exercise wisdom.

In summation, it can be argued that multiple-choice examinations, even those consisting primarily of higher-order questions, provide only a superficial assessment of a neurosurgeon’s expertise in patient management, with limited representation of a neurosurgeon’s intuition and decision-making. Accordingly, oral board exam failures are frequently attributed to inappropriate surgical indications and subtle errors in judgment, rather than a lack of factual knowledge. Therefore, it is essential to further evaluate the performance of LLMs in this domain, which will be the subject of future study. As AI continues to advance, multiple-choice examinations may assume a less prominent role in medical education, with oral boards-style verbal assessments gaining increased importance. Another change that AI may bring to neurosurgical education is the use of LLMs by neurosurgical trainees for boards preparation. For example, with the initial input of a clinical scenario to discuss, an LLM like GPT-4 may be used as a conversational aid to rehearse the discussion of more challenging topics for the boards or even appreciate new clinical insights or rationales from the responses generated by LLMs.

While this multiple-choice question bank cannot fully capture the dynamic, conversation-based, and open-ended nature of the oral boards, our findings do hint at the potential value of LLMs such as GPT-4 in neurosurgical education and in clinical decision-making. Given a score improvement of over 20% between two AI models released just four months apart, it is critical for neurosurgeons to stay informed and up-to-date about these rapidly evolving tools and their potential applications to clinical practice. Toward this end, the development of methods to quantify and understand hallucinations, as well as the validation of LLMs on higher-order and open-ended scenarios, is vital for the successful integration of these tools into neurosurgery and other high-stakes medical specialties. Ultimately, the capacity and extent to which LLM’s are incorporated into practice will depend heavily on the ability to minimize and recognize hallucinations. In summary, this study represents an important initial benchmark in evaluating LLM performance in higher-order and relatively more open-ended clinical scenarios.

### Limitations

This study has several potential imitations. First, as discussed earlier, the use of multiple-choice to quantify LLM knowledge for higher-order neurosurgical topics incompletely captures the open-ended nature of the true neurosurgery oral board examination. We aim to conduct more open-ended assessments of LLM neurosurgical knowledge in future assessments. Second, it is possible that certain question characteristics, such as incorporation of higher-order problem-solving, may have been characterized incorrectly or differently by a separate evaluator. However, this study utilized three authors to collect data on question characteristics and differences between evaluators was minimal (<5%). Third, the accuracy of LLMs when incorporating imaging-related data could not be assessed due to these functions not being presently being publicly available, and LLM performance on imaging requests following multimodal input will be the subject of future study. Fourth, due to continual “under the hood” improvements to LLMs influenced by factors like aggregate user input, the performance of LLMs like GPT-4 may change gradually and returned answers may differ from how they are presented in the current study. In order to minimize heterogeneity due to these factors, data collection was performed in a 24-hour range for each separate LLM.

## Conclusion

On a question bank of predominantly higher-order management case scenarios intended for neurosurgery oral boards preparation, GPT-4 achieved a score of 82.6%, outperforming GPT-3.5 and Google Bard. Unlike GPT-3.5 and Bard, higher-order problem-solving was not predictive of lower answer accuracy from GPT-4. Finally, GPT-4 exhibited significantly lower rates of hallucinations on imaging-related questions.

## Data Availability

All data produced in the present study are available upon reasonable request to the authors.

https://github.com/oliverytang/chatgpt_neurosurgery

## Notes

**Conflicts of Interest:** The authors report no conflict of interest concerning the materials or methods used in this study or the findings specified in this paper. However, we would like to acknowledge and thank the Congress of Neurological Surgeons for their development and dissemination of the mock exam questions used for this study.

### Competing Interest Statement

The authors have declared no competing interest.

### Funding Statement

This study did not receive any funding.

